# Signal Intensity Normalization of Multispectal Magnetic Resonance Images Obtained with Clinical Sequences

**DOI:** 10.1101/2021.10.27.21265570

**Authors:** Matthew R. Brier, Biao Xiang, Zhuocheng Li, Robert T. Naismith, Dmitriy Yablonskiy, Anne H. Cross, Tammie L. S. Benzinger, Abraham Z. Snyder

**Affiliations:** Department of Neurology, Washington University in St. Louis; Department of Radiology, Washington University in St. Louis

## Abstract

Assessment of intrinsic tissue integrity is commonly accomplished via quantitative relaxometry or other specialized imaging, which requires sequences and analysis procedures not routinely available in clinical settings. We detail an alternative technique for extraction of quantitative tissue biomarkers based on intensity normalization of T1- and T2-weighted images. We develop the theoretical underpinnings of this approach and demonstrate its utility in imaging of multiple sclerosis.

## 1. Introduction

Magnetic resonance imaging (MRI) is the premier tool for diagnosis and monitoring of numerous neurological conditions [5, 22, 10, 29]. Focal lesions (e.g., multiple sclerosis [MS] plaques, brain tumors, infarcts) are readily detectable both visually and through automated segmentation algorithms [4, 12, 30, 32]. However, additional subtle abnormalities are increasingly recognized as important [23, 1, 21] in MS [3] as well as other conditions [15]. Conventionally, quantification of abnormalities in normal appearing tissue requires specialized sequences or extensive image processing [20, 16, 27], which generally precludes application in routine clinical practice. In this work, we characterize a recently described technique [3] for the estimation of standardized intensity properties in images obtained with clinically available T1-weighted and T2-weighted (FLAIR) sequences.

T1 and T2 are intrinsic properties of tissue that depend on the underlying cellular composition [28, 35]. Reconstructed intensities in T1- and T2-weighted images depend on the intrinsic properties of the tissue being studied interacting with sequence parameters, spatial encoding radiofrequency (RF) field (B1), and the main field (B0) [2]. However, reconstructed image intensities also vary with uncontrolled factors including quality of shim, head size, temperature, etc. These dependencies preclude direct, numerical comparison of T1- and T2-weighted intensities across sessions or individuals. Standardization of T1- and T2-weighted image intensities would facilitate comparison across sessions or individuals and allow for new questions to be asked in legacy or clinical imaging data.

We previously reported results obtained by standardization of T1-weighted (T1w) and T2-weighted (T2w) images using an intensity histogram normalization procedure applied to data from a multinational collaboration (Multiple Sclerosis Partners Advancing Technology and Health Solutions; MS PATHS) [3]. In this procedure, following standard image preprocessing (e.g., spatial alignment, skull stripping), bivariate voxel-intensity histograms are constructed from the T1w and FLAIR (T2w) data. These histograms are affine registered to a standard intensity histogram template, thereby standardizing intensity values across subjects. In our previous work, we obtained signal mean and standard deviation estimates for each image (T1w, FLAIR) and tissue class (cerebrospinal fluid, gray matter, white matter; CSF, GM, WM) by decomposing intensity normalized histograms using Gaussian mixture modeling (GMM). When evaluated in cross-sectional data from patients with MS, these GMM features separated MS patients from controls, distinguished between MS subtypes, and correlated more closely with MS-related disability than more conventional lesion-based imaging measures. Having demonstrated the utility of the intensity normalization approach, we now demonstrate its applicability to longitudinal datasets. We first describe the intensity normalization algorithm in greater detail and then demonstrate test-retest reliability of derived measures. More specifically, we describe reliability of lesion segmentation as well as intensity-based measures within *a priori* regions of interest.

## 2. Theory

Objectives of imaging in the present context include distinguishing between tissue classes (e.g., MS lesions vs. normal appearing WM) and quantification of abnormalities in those tissues (e.g., WM in controls vs. patients). It has been established that this can be done on the basis of quantitative determination of T1, T2, and related quantities [20, 35, 31]. But, as discussed above, quantitative measurement of T1 and T2 cannot be reliably done on the basis of T1w and T2w images.

However, in theory, there exists an informational equivalence between T1 and T2 vs. T1w and T2w images can be formally cast in terms of Kullback-Liebler divergence, *I*(1:2). At the single voxel level, *I*(1:2) is information in favor of *H*_1_ (e.g., lesion) vs. *H*_2_ (e.g., normal appearing WM)^1^ [17]. To further consider this issue theoretically, define *X* = {*T*1*w, FLAIR*}_*norm*_ (normalized intensity measure) vs. *Y* = {*T*1, *T*2} (absolute measures). The question, then, is *I*(1:2; *X*) comparable to *I*(1:2, *Y*)? Invariance theory states that *I*(1:2, *X*) ≤ *I*(1:2; *Y*) with equality if there exists a smooth, invertible relation between *X* and *Y*. Thus, *I*(1:2;*X*) = *I*(1:2; *Y*) if there exists some relation, *F*, such that *Y* = *F*(*X*) and *X* = *F* ^−1^(*Y*). The algebraic proof is remarkably simple when *X* and *Y* are multivariate Gaussian distributions and *F* is a linear transform (see p. 194 of [17]). However, it is not necessary that *F* be linear (or affine), only that it be non-singular (see Fig. 5 in [31]). This condition presumably applies to the relation between {*T*1*w, FLAIR*}_*norm*_ and {*T*1, *T*2}, as it may be assumed that reconstructed image intensities depend smoothly on the physical properties of brain tissue and variation in MRI pulse sequence parameters. We empirically demonstrate this principle below (Section 4.1).

We have so far considered discrimination between tissue classes at the single voxel level. We now extend the theory to the whole brain. As was noted above, the information in {*T*1*w, FLAIR*}_*norm*_ and {*T*1, *T*2} is equivalent provided that there exists a smooth relation, *F*, connecting the two measures. Although it may be assumed that a smooth *F* exists at every voxel, this relation may not be the same everywhere in the brain. Nevertheless, we here empirically assume a fixed relation between {*T*1*w, FLAIR*}_*norm*_ and {*T*1, *T*2}. Our hypothesis is based on the observation that intensity normalization, described below, can mitigated multiple sources of variance including magnetic field inhomogeneities. Accordingly, the experimental question becomes whether this approach supports reliable tissue characterization. Thus, we propose to compensate for uncontrolled factors that influence T1w and T2w image intensities by requiring that {*T*1*w, FLAIR*} intensity histograms conform to a standard intensity template. At issue, then, is the extent to which normalized intensity histograms of the form described below (Section 3.3) can be used to characterize tissue in place of quantitatively measured T1 and T2.

## 3. Materials and Methods

### 3.1. Study Design, Participants, and Image Acquisition

This study makes use of two datasets. The first dataset included *N*_1_ = 7 healthy young adults with both quantitative T1 imaging as well as clinical-type T1w images. This dataset was used to demonstrate the theoretical informational equivalence between weighted and absolute intensity histograms. The second dataset included *N*_2_ = 30 subjects with MS who were repeatedly studied using clinically available, high resolution T1w (MP-RAGE) and FLAIR (T2w) sequences. This dataset was used to demonstrate the test-retest reliability of intensity normalized measures. Each of these datasets are small compared to existing datasets but benefit from containing specific sequences (e.g., quantitative T1 imaging) or are densely sampled (e.g., 4 imaging sessions within the span of a week) and are thus suitable for technique development.

#### 3.1.1. Quantitative vs. Weighted Comparison Cohort

Seven healthy controls (2 male, 5 female, age range 22–27 years) were imaged for the comparison of the informational content of intensity normalized clinical images vs. absolute T1 estimation. All participants were neurologically normal, able to undergo MRI, and provided informed consent. Imaging was conduced on a 3T Prisma scanner (Siemens, Erlangen, Germany) equipped with a 32-channel phased-array head coil. T1w images were acquired with an MP-RAGE sequence (0.8mm isotropic voxels, TR 2.4s, TE 2.22ms, inversion time 1s, *α* = 8°). For quantitative T1 mapping, voxels were 1.3mm isotropic, acquired using three-dimensional multi-gradient-echo sequence with five flip angles (*α* = 5°, 10°, 20°, 40°, 60°) and three gradient echo times (*TE* = 3*ms*, 7*ms*, 11*ms*) for each *α*. The sequence incorporated a generalized auto-calibrating, partially parallel acquisition algorithm [13] with an acceleration factor of 2, and 24 auto-calibrating lines in each phase encoding direction. T1 maps were generated by fitting the combined data to the Ernst equation [9] with *B*1 correction [36] that accounted for the imperfect radio frequency spoiling and transverse magnetization relaxation.

#### 3.1.2. Test-Retest Reliability Cohort

Thirty MS patients (17 female, age ranged 21–55 years, EDSS [18] 0–6) were enrolled in the reliability sub-study of MS PATHS [24]. MS PATHS is a natural history study funded by Biogen in which participating centers contribute standardized imaging and behavioral data from patients with MS. For the reliability sub-study, each participant underwent four MRI sessions on two scanners over two days. Scanning sessions on the same day were 1–6 hours apart and scanning days were 2–7 days apart. Thus, each participant contributed 4 repeated MP-RAGE (T1w; field of view 256mm × 256mm, 1mm isotropic voxels, TR 2.3s, TE 2.98ms, TI 900ms) and 4 repeated FLAIR (T2w; field of view 256mm × 256mm, 1mm isotropic voxels, TR 5s, TE 393ms, TI 1800ms) images of the same brain^2^. All imaging was acquired on Siemens 3T scanners; model of scanner varied with acquisition site.

### 3.2. Image Pre-processing

T1w and FLAIR images from each session were 9-parameter affine coregistered (rigid body + cardinal axis scaling) and the data obtained in all four sessions were mutually coregistered using in-house software^3^. Following composition of transforms, all images were resampled (one step) in a standard atlas space (1mm^3^ voxels). Images then underwent brain extraction (BET) [33] and bias field correction (FAST) [37], implemented in the FSL toolbox [14]. Intensity values in each weighted image were scaled (one multiplicative constant per image) to obtain a voxel intensity distribution mode value of 1,000 over the whole-brain; this scaling accelerates subsequent intensity normalization.

### 3.3. Image Intensity Histogram Normalization

The goal of image intensity histogram normalization is the standardization of bivariate intensity (T1w and FLAIR) distributions to match a standard template [3]. Prior to normalization, histogram features are grossly similar across individuals but vary in extent and skew owing to factors of non-interest (e.g., proximity of the head to the receiver coils) in addition to biologically relevant factors (e.g., disease status). To accomplish normalization, each individual’s data were 6-parameter affine transformed to match the template. Histogram similarity was evaluated as the mean-squared error between the template and transformed individual histogram. Optimization was achieved by regular step gradient descent [34] implemented in MATLAB 2020a (Mathworks, Natick, MA). Let *A* be the affine transform that minimizes the difference between each individual’s data and the template. This matrix has the form

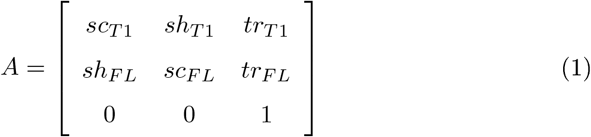

where *sc, sh*, and *tr* represent scaling, shearing, and translation along either the T1w or FLAIR axis. Let *X* = {*T*1*w, FLAIR*} represent an individual’s bivariate intensity histogram prior to normalization. Multiplication of *X* by optimized *A*

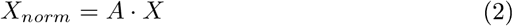

yields the normalized bivariate intensity histogram. Voxelwise application of *A* to the bivariate data generates a T1w and FLAIR images that have been intensity normalized to a standard template.

### 3.4. Image Tissue Scores and Gradients

Tissue classes are represented as distributions around centroids in bivariate intensity space. Tissue class centroids were defined *a priori* by manual segmentation of a single representative image. For each voxel, a continuous score *u*_*i*_ ∈ [0, 1] was assigned for each tissue class *i* such that ∑ _*i*_ *u*_*i*_ = 1. The algebra regarding the calculation of *u*_*i*_ follows.

For any multispectral image voxel, the standardized squared distance from the *i*-th tissue class centroid 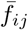 is given as

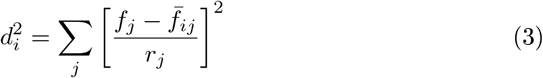

where *j* indexes contrasts (e.g., T1w, FLAIR) and *r*_*j*_ is the intensity range containing non-artifactual intensity values of contrast *j*. Raw membership in class *i* varies as 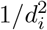 and the normalized membership class as

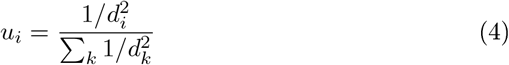

where *k* is a dummy summation index. Tissue class membership can be defined by identifying the class *i* with the largest *u*_*i*_ (‘winner-take-all’ [WTA] segmentation).

Tissue class gradients have multiple uses in the analysis of multi-spectral images. Let *D*_*j*_ ≡ *∂/∂f*_*j*_. It follows,

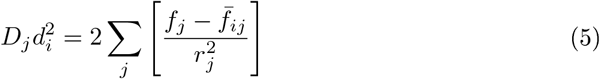

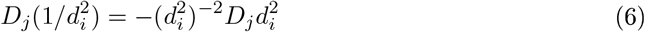

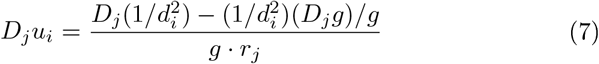

where 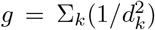 and 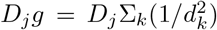. Application of the chain rule yields tissue class membership gradients in image space

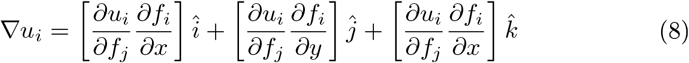

where 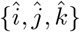 are the (*x, y, z*) cardinal unit vectors in image space.

Boundaries between tissue compartments correspond to loci at which two class memberships are dominant and approximately equal. In the archetypical case, *u*_1_ = *u*_2_ = 0.5, taking classes 1 and 2 as examples. In general, the tissue class boundary corresponds to *u*_1_ ≈ *u*_2_, with *u*_1_ + *u*_2_ < 1, because of finite representation of other tissue classes (∑ _*i*_ *u*_*i*_ = 1). Figure 1 illustrates a toy case in which the tissue class boundary normal points exactly in the *x*-direction. The boundary occurs where |*u*_2_(*du*_1_*/dx*) − *u*_1_(*du*_2_*/dx*)| is maximal. In 3-D, this expression generalizes to the locus where |*u*_2_∇*u*_1_ − *u*_1_∇*u*_2_| is maximal. A rationale for this expresion may be seen in the equality

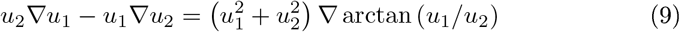

**Figure 1:**
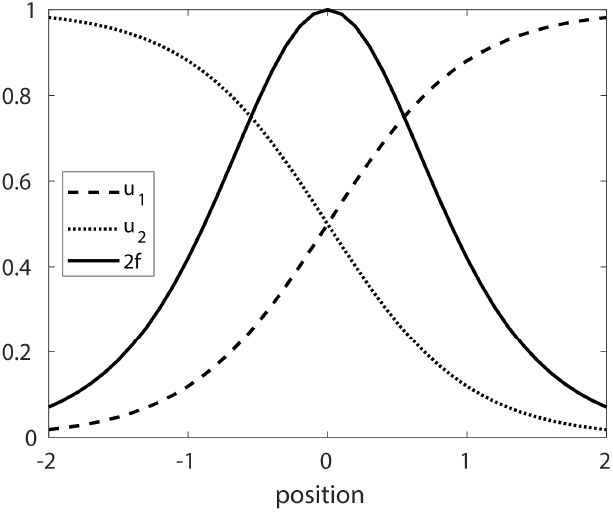
Toy case illustrating the tissue class boundary condition. Let *u*_1_ = *e*^*x*^*/* (*e*^*x*^ + *e*^−*x*^) and *u*_2_ = *e*^−*x*^/(*e*^*x*^ + *e*^−*x*^). Trivially, *u*_1_ + *u*_2_ = 1 (in this case). Evaluating |*u*_2_ (*du*_1_*/dx*) *− u*_1_ (*du*_2_*/dx*)|, we obtain 2*/* (*e*^*x*^ + *e*^−*x*^) ^2^ = *f*. The algebra in this toy case has been simplified by the use of logistic functions. However, Equation 9 applies wherever there exist relatively sharp transitions between low vs. high tissue class values.

The term ∇ arctan (*u*_1_*/u*_2_) attains a maximal value when *u*_1_ = *u*_2_. Thus, a boundary corresponds to a locus at which *u*_1_ ≈ *u*_2_ and 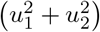 is substantial.

## 4. Results

### 4.1. Demonstration of Informational Equivalence Between Weighted and Absolute Intensity Distributions

Section 2 established the theoretical equivalence of two multivariate histograms provided that they are related via a non-singular (invertible) transformation. Here, we empirically demonstrate the existence of such a transform between reconstructed intensities in a T1w image and quantitative T1. Figure 2 shows the T1w and T1 histograms of a representative subject. Note inversion of the horizintal axes (*T*1 ∝ 1*/T*1*w*). Nevertheless, comparable features are evident: two prominent peaks corresponding to GM and WM and a long tail at small values (in T1w) or large values (in T1) representing CSF. Thus, it is intuitively plausible that the histograms contain similar information.

**Figure 2:**
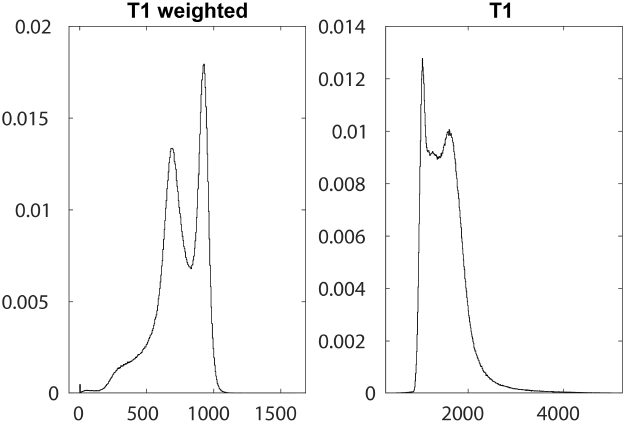
T1w and T1 intensity histograms in a representative subject. Image intensities were extracted from voxels within the brain mask and displayed as histograms. Note inverted order of histogram features: In the T1w image, tissue classes are ordered CSF < GM < WM. In contrast, measured T1 values are ordered WM < GM < CSF.

We demonstrate that T1w and T1 values can be made equivalent by histogram matching. Let *C*_*T*1_ and *C*_*T*1*w*_ be the cumulative density function of the image intensity histograms corresponding to the T1 and T1w images. The transformation between the two histograms therefore is

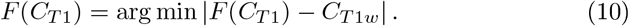

The resulting transformed histograms are shown as cumulative density functions in Figure 3. The T1w function (black solid line) and the transformed T1 function (green dashed line) are nearly identical. Note that the mapping (*F*) is different in each subject. We evaluated error as *ϵ*^2^ = ∑ (*C*_*T* 1*w*_ − *Ĉ*_*T*1_)^2^ where *Ĉ*_*T*1_ is the cumulative density function with *F* applied. Mean *ϵ*^2^ across subjects was 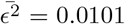. This result demonstrates the existence of a mapping between the intensity histograms of T1w and absolute T1 images. Figure 3 demonstrates 1-D histogram matching. In principle, 2-D histogram matching could be similarly done *in normal participants*, provided that each had the same proportion of tissue classes. However, this is not possible in MS patients as the proportion of tissue classes, especially lesion, varies greatly. In the following, we report results obtained by affine transformation of 2-D histograms.

**Figure 3:**
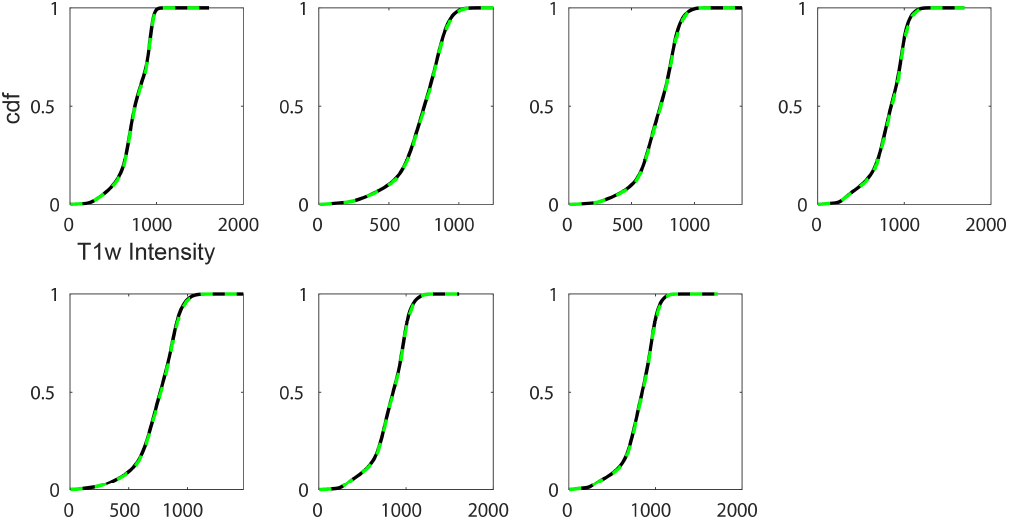
Informational equivalence between T1 and T1w images. Superimposed, individual subject (*N*_1_ = 7), T1w cumulative density (black) and transformed T1 cumulative density functions (dashed green). Note near identity of the two cumulative density functions. Thus, there exists a [generally different] *F* for each subject.

### 4.2. Intensity Normalization

Figure 4 illustrates 2-D histogram matching by affine transformation (Eq. 2) in two example subjects. The first panel shows the template intensity histogram. The top row shows the template and individual subject histograms overlayed. Following intensity normalization (bottom row), the overlap between the individual subject histograms and the template is markedly improved. Alignment is determined by minimizing the mean squared error between the template and the aligned image.

**Figure 4:**
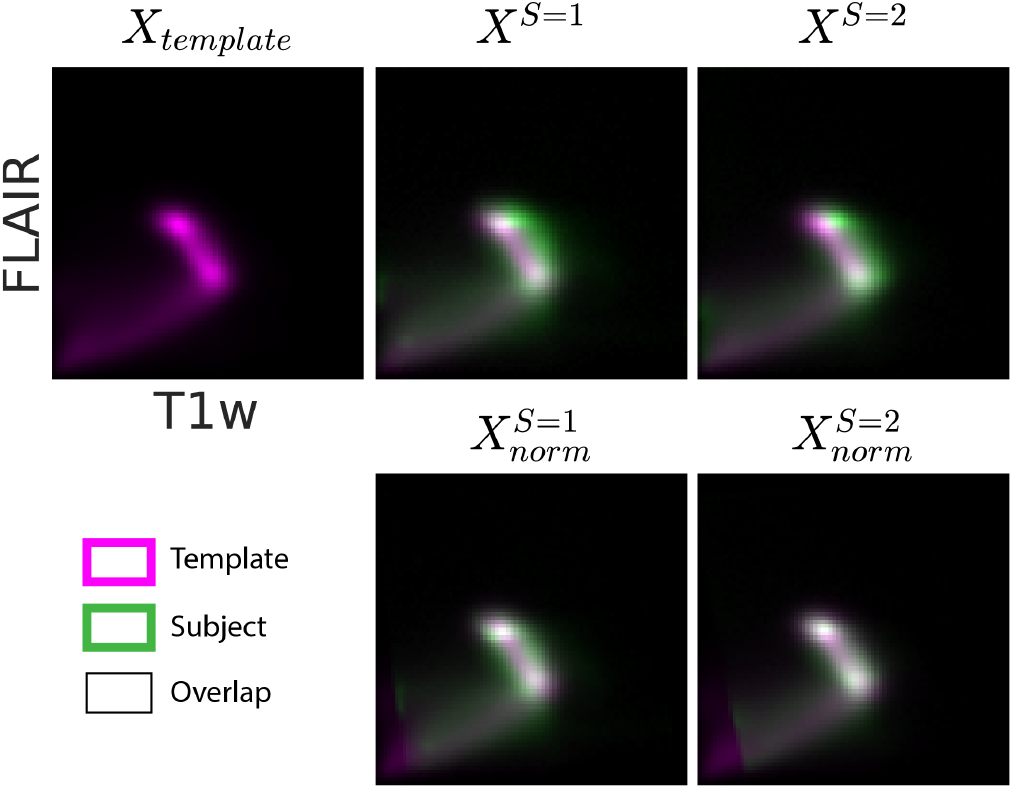
Bivariate intensity histogram normalization improves the match between single-subject data and the template. The top left panel shows the template bivariate intensity histogram, *X*_*template*_, in purple. The horizontal and vertical axes index T1w and FLAIR (T2w) intensities, respectively. The two panels in the top row show the template histogram with “raw” single subject histograms overlayed in green. The union overlap of green and purple is displayed as white. The bottom rows show the results of intensity normalization using Equation 2. Objective function error before and after affine transformation was 24.7 → 11.5 and 30.8 → 10.7, respectively, for the two subjects.

**Figure 5:**
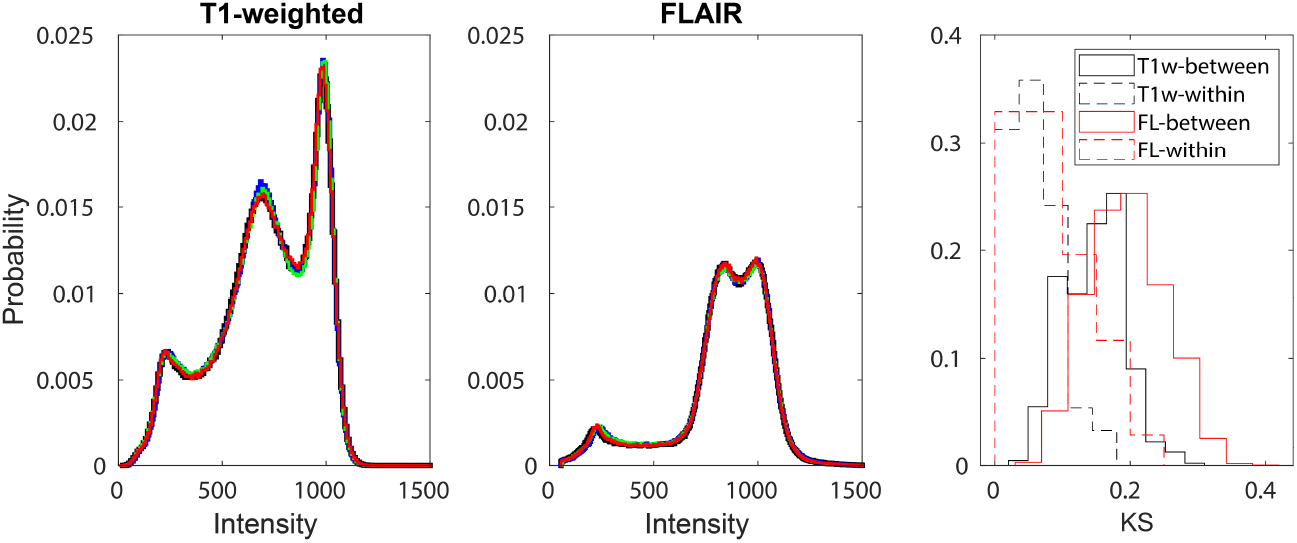
Normalized image intensity histograms are reliable across imaging sessions. Intensity normalized image intensity histograms from a single representative subject are shown for the T1-weighted and FLAIR images. The four test-retest sessions are shown in different colors. The critical feature is nearly perfect overlap across sessions. The rightmost panel shows the distribution of K-S statistics for within-subject and between-subject histogram comparisons for T1-weighted and FLAIR.

### 4.3. Within-Subject Reliability of Intensity Normalization

Intensity histogram normalization generates T1w and FLAIR images with standardized intensities. Figure 5 addresses within-subject and cross-subject comparison of normalized intensity histograms. It may be assumed that there was no interval change over sessions in these subjects. The left two panels show normalized T1w (left) and FLAIR (middle) image intensity histograms for a representative subject superimposed over the four sessions. Each scanning session is represented by a different color. The salient feature of these histograms is their striking consistency.

We quantified histogram similarity using the two-sample Kolmogorov–Smirnov (K-S) test statistic, *KS*_*nm*_ = sup_*x*_ |*C*_1,*n*_(*x*) − *C*_2,*m*_(*x*)|, where *C*_1,*n*_ and *C*_2,*m*_ are the two probability distributions. Small values of *KS* indicate similar distributions. We calculated the K-S statistic between all histogram pairs (4×30 = 120) within a contrast mechanism (T1w, FLAIR), distinguishing between within-subject vs. between-subject comparisons. The distributions of these K-S values are shown in the right panel of Figure 5. Note small K-S statistics representing within-subject comparisons for both T1w and FLAIR images. These results provide further empirical evidence that the intensity normalization procedure is reliable. Generally larger cross-subject K-S values reflect individual differences.

We next evaluated test-retest reliability tissue class scores within GM, WM and lesion, determined by WTA classification of intensity-normalized images. Mean (*μ*) and within-tisue class standard deviation (*σ*) signal intensities were calculated for T1w and FLAIR images, for all sessions and subjects. Reliability was quantified using intra-class correlation (ICC) for T1w *μ*, FLAIR *μ*, T1w *σ*, and FLAIR *σ* separately. This calculation was performed with sessions as repeated measures and subjects as the ensemble average. Resulting ICC values are shown in Table 1. Large ICC values indicate reliable tissue score mean and variability estimates.

**Table 1:** ICC representing the reproducibility of tissue characteristic estimates over sessions, averaged across subjects.

| | T1w $\mu$ | FLAIR $\mu$ | T1 $\sigma$ | FLAIR $\sigma$ |
| --- | --- | --- | --- | --- |
| WM | 0.81 | 0.80 | 0.89 | 0.93 |
| GM | 0.89 | 0.90 | 0.95 | 0.80 |
| Lesion | 0.89 | 0.95 | 0.75 | 0.86 |

We repeated the tissue class score ICC analysis in large, subcortical structures segmented in the T1w image using the FIRST tool implemented in FSL [26]. Note generally high ICC vlaues (*>* 0.7) in these structures (Table 2).

**Table 2:**
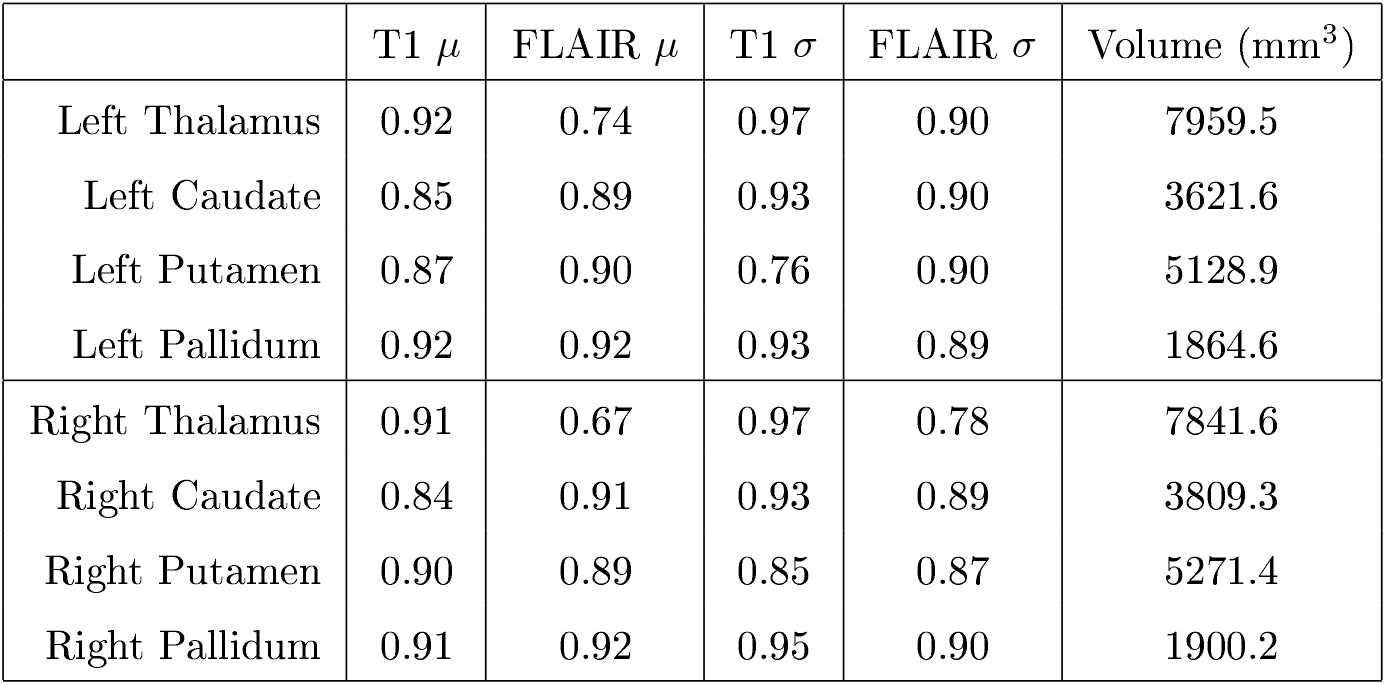
ICC for image intensity parameters within subjects across sessions. Mean structure volume is also shown.

### 4.4. Between-Subject Reliability of Intensity Normalization

We next assess across-subject reliability of 2-D histogram normaliztion in the reproducibility cohort. This assessment is complicated by substantial variation in the degree of MS severity. Hence, some variability of intensity normalized histograms is expected. Nevertheless, major features of these histograms should be similar. The first column of Figure 6 shows the mean T1w and FLAIR intensity normalized histogram (black line) with the single subject histograms (green lines) superimposed. To further clarify the reliability across subjects, the second and third columns of Figure 6 show the peaks corresponding to the GM and WM locus. Peak locations vary across subjects but there is a strong central tendancy. The box plots in the final column show how the peak location varies across subjects. This display demonstrates that histogram normalization produces comparable intensity histograms across subjects.

**Figure 6:**
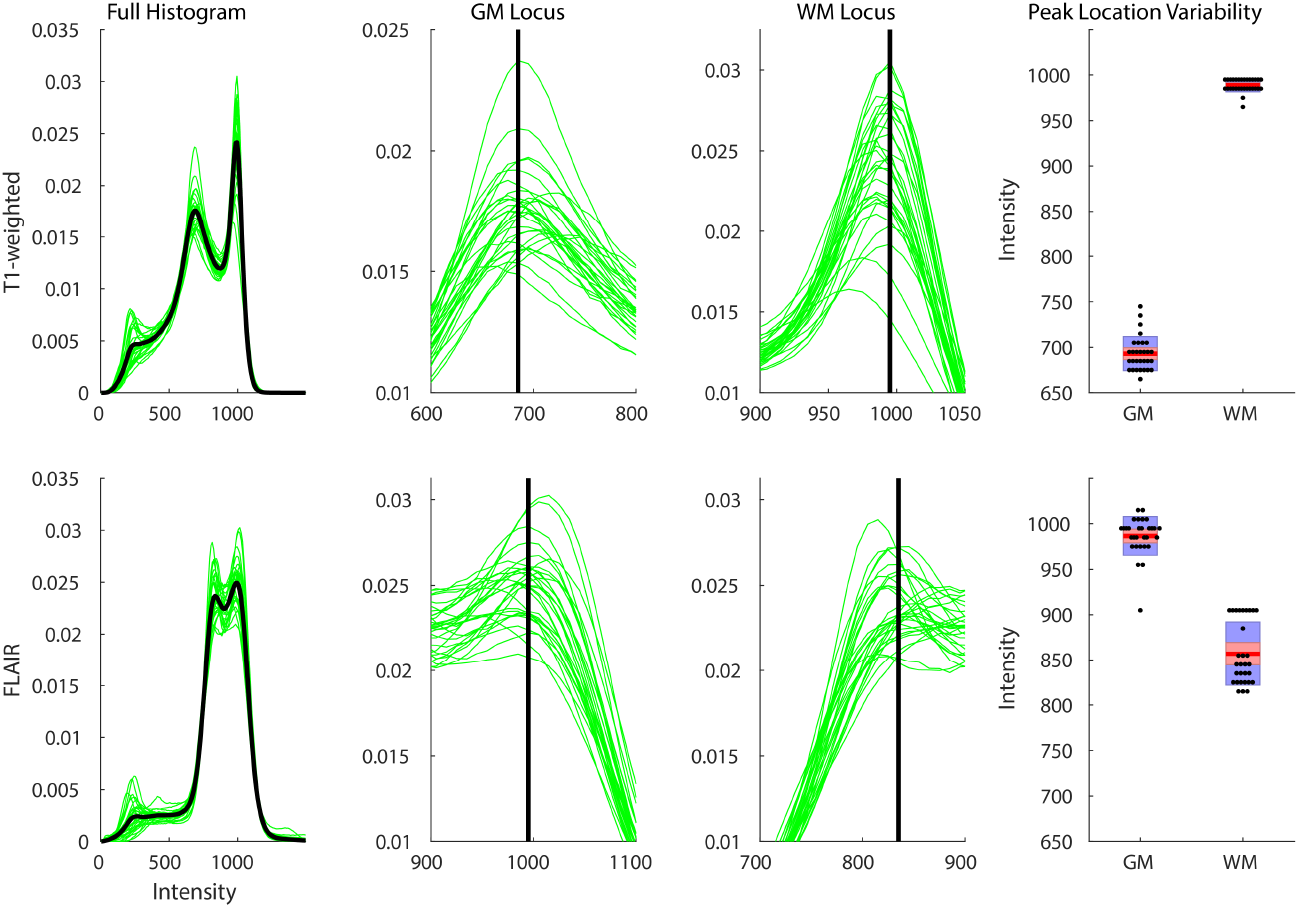
Intensity Normalization Across Subjects. The top and bottom rows show T1w data and FLAIR data, respectively. The first column shows the mean (across subjects) intensity histogram (black lines) with individual subjects superimposed (green lines). The second and third columns are restricted to the region around the GM and WM loci. Vertical black lines indicate locus center-of-mass. The fourth column shows the distribution of peak location across subjects.

### 4.5. Tissue-Score and Gradient-Based Lesion Segmentation

Image segmentation can be accomplished by labeling each voxel according to the tissue class, *i*, with the greatest value of *u*_*i*_ (WTA approach). Although this is possible for all tissue classes, we focus here on lesion segmentation. Figure 7 shows the “raw” WTA segmentation result in a representative MS patient. Note the presence of voxels surrounding lesions, misclassified as GM (Figure 7). This misclassification is attributable to image noise and volume averaging interacting with proximity of the GM centroid to the trajectory connecting the centroids corresponding to lesion and WM (Figure 7). We describe a novel approach to solving this problem incorporating tissue score gradients, ∇*u*_*i*_.

**Figure 7:**
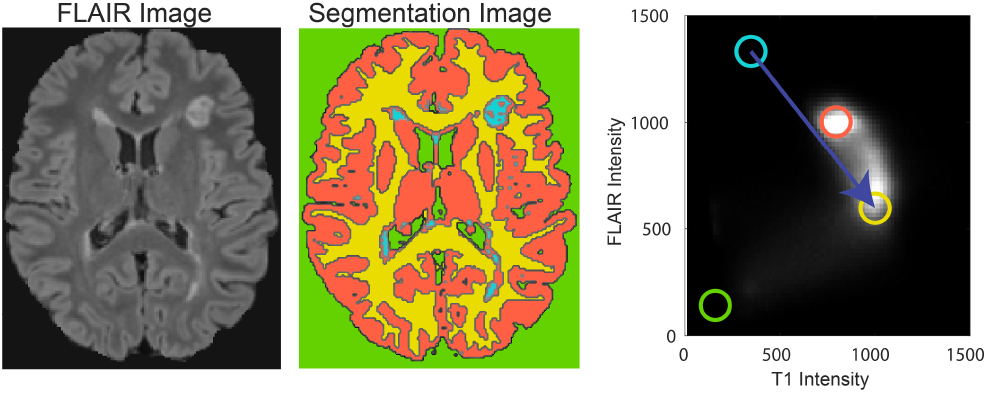
Tissue-Score Based Segmentation. The left panel shows a FLAIR image from a representative MS patient. The WTA segmentation, shown in the middle panel, plausibly corresponds to the FLAIR image. CSF is green; WM is yellow; GM is orange; lesion is blue. Note the presence of voxels misclassified as GM surrounding lesions. A heuristic explanation for this misclassification is illustrated in the right panel: The GM centroid (orange circle) is close to the trajectory (dark blue arrow) connecting the lesion centroid (light blue circle) and the WM centroid (yellow circle).

Figure 8 illustrates one FLAIR slice containing a prominent MS lesion. The lesion as well as a lesion-free region of brain are shown in close-up. Tissue class gradients (∇*u*_*i*_) are represented as quiver plots. Note prominent true WM and true lesion gradients as well prominent factitious GM gradients surrounding the lesion. Note absence of lesion gradients in normal appearing WM and presence of lesion gradients inside the lesion boundary. The key feature that enables correction of GM misclassification at the lesion boundary is ∇*u*_*GM*_ ≈ −∇*u*_*Les*_; hence, ∇*u*_*GM*_ · ∇*u*_*Les*_ is strongly negative. Identification of this feature enables correction of the misclassification (see Fig. 9).

**Figure 8.**
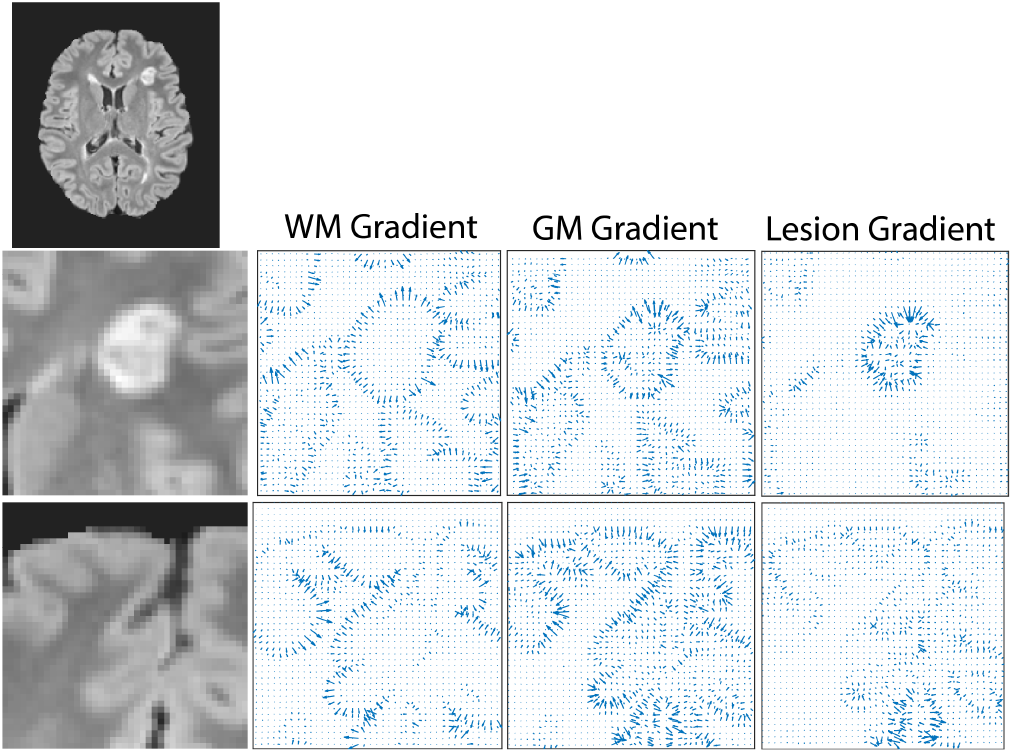
Tissue Class Gradients. The top panel shows one slice of a FLAIR image from a representative MS patient. Two regions are shown in close up in lower rows. The first region contains a stereotypical lesion; the second region is lesion-free. Tissue score gradients corresponding to WM, GM, and lesion are shown as ‘quiver’ plots in which length encodes gradient magnitude. Arrowheads point towards increasing tissue class score. Note true WM gradients and lesion gradients as well as factitious GM gradients surrouding the lesion.

**Figure 9:**
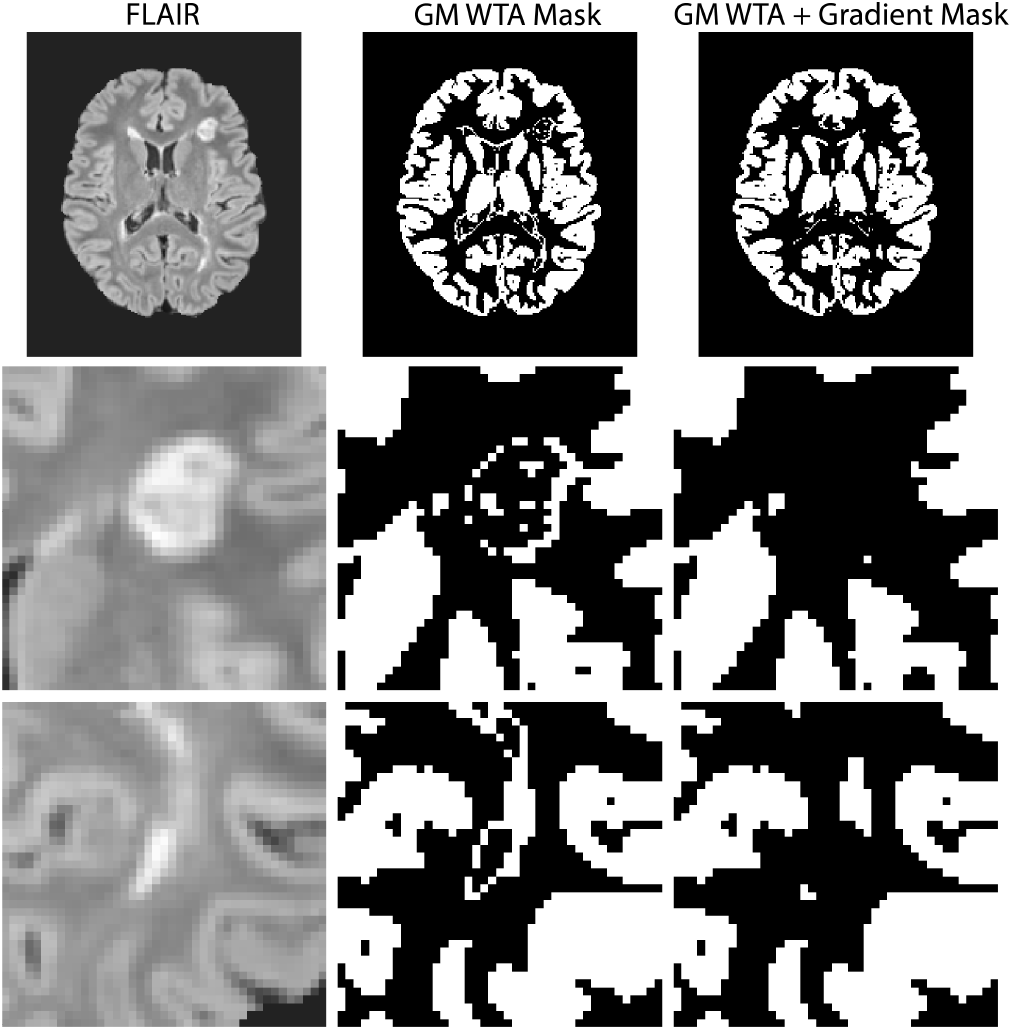
Correction of factitiously labeled GM voxels using tissue score gradients. The left columns shows one slice of a stereotypical FLAIR image from a MS patient. Two regions are shown in close-up in the second and third rows. The second column shows voxels classified as GM prior to correction of misclassification. The third column shows voxels classified as GM following gradient-based correction at lesion boundaries. Note substantial mitigation of misclassification.

Figure 9 shows a FLAIR slice with several prominent lesions shown in closeup. The second column shows voxels classified as GM, demonstrating misclassification surrounding lesions. This misclassification was corrected using an emprirical rule: GM voxels within a radius of 2mm of any lesion voxel, in which ∇*u*_*GM*_ · ∇*u*_*Les*_ < 0.1, were assigned to the next most highly scored tissue class. This procedure greatly reduced the factitious ring of misclassified GM surrounding lesions (Figure 9).

Finally, we examined the reliability of lesion segmentation using the presently described lesion segmentation technique (Section 4.5). Measured total lesion volume in the brain and the measured individual lesion volume should not change across the four imaging sessions. To avoid spurious identification of lesion voxels at the brain edge (e.g., as illustrated in Fig. 8) this analysis was restricted voxels identified as WM by FAST segmentation of our standard MP-RAGE atlas template [14]. Figure 10 shows the correlation between total lesion burden pairwise for all session combinations within a subject for all subjects. In total, there are 30 · _4_*C*_2_ points represented. The agreement across imaging sessions is very good (*r* = 0.97). Similarly, we identified spatially coincident lesions in each session and compared the segmented volume across sessions. This correlation also is very strong (*r* = 0.99). Thus, our lesion segmentation algorithm produces stable lesion extent estimates. Lastly, we compared the segmentation results derived from our technique and an existing algorithm in the literature [6] for all 30 · 4 imaging sessions. The correlation in recovered total lesion volume was high (*r* = 0.94) and the correlation between individual lesion volumes was similarly high (*r* = 0.95). One note is that the slope of the total lesion volume scatter plot (Figure 10, left-lower panel) is not 1, indicating small differences in lesion boundary sensitivity and specificity. Overall, this supports the reliability and validity of the present approach.

**Figure 10:**
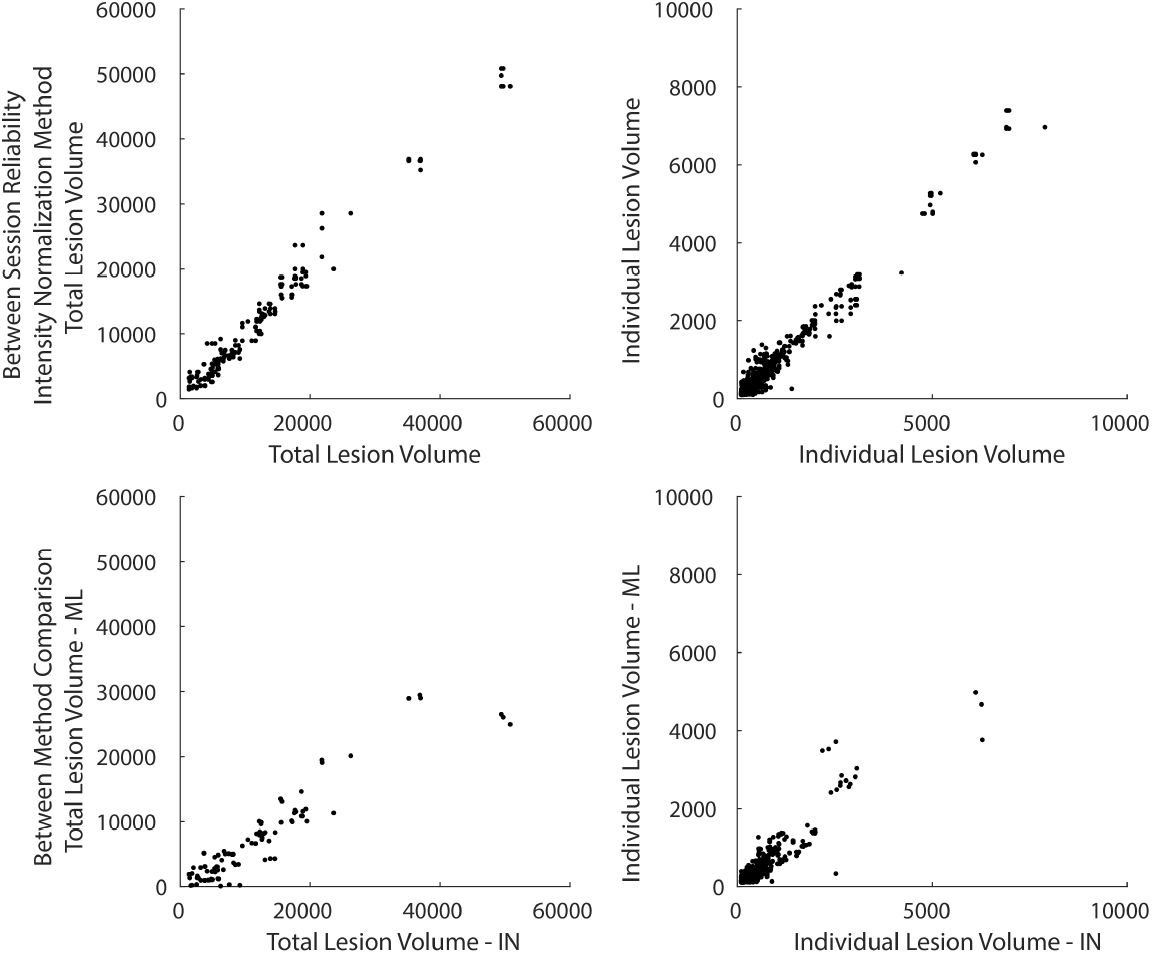
Segmented lesion volume is relable across scanning sessions. The upper-left panel shows the correlation between total lesion burden across session pairs within subject, compiled over all subjects using the intensity normalization approach. The upper-right panel shows the correlation between individual lesion volumes across session pairs (within subject) compiled over all subjects using the intensity normalization approach. In both cases, the correlation is very strong, indicating reliable lesion segmentation using the procedure illustrated in Figure 9. The bottom panels display the same information comparing the intensity normalization approach with an existing approach in the literature [6]. The correlation between the two methods is high indicating similar lesion segmentation. IN - Intensity normalization approach, ML - machine learning approach described in [6]

## 5. Discussion

Extraction of voxelwise biomarkers of pathology using quantitative relaxometry has been well established [28, 20]. Here, we present an alternative technique which uses intensity normalization of data acquired using standard FDA-approved sequences. Basic information theoretic considerations predict that this is possible; we demonstrate this principle using histogram matching (Figure 3). We show that the present approach, based on affine transformation of bispectral histograms (Figure 4), generates reproducible, quantitative biomarkers of tissue properties. Additionally, these voxelwise measurements facilitate tissue segmentation with minimal dependence on template priors. The present technique generates continuous tissue class membership scores. Spatial gradients of these scores are useful for refining tissue segmentation and potentially could be useful in longitudinal studies of subtle changes in lesion boundaries and within-lesion heterogeneity.

We began by claiming that two histograms *A* and *B* are informationally equivalent if there exists a transform *A* = *F* (*B*) [17]. *F* need not be linear (or affine) and only must be smooth and invertible. When applied to T1w images with a known corresponding T1 image, *F* can be attained by histogram matching. However, when applied to images absent a known underlying distribution, direct histogram matching is not feasible given the unknown target. To circumvent this limitation, we utilize affine registration of intensity histograms to a template derived from normal control data. Affine registration allows for the preserved representation of biologically relevant features in the registered data. Such features include discrete lesions or tissue atrophy in addition to changes in intrinsic tissue intensity. As implemented here, affine registration of bispectral intensity histograms reduces the contribution of factors of no interest (e.g., differences in scanner characteristics, head coil differences) while allowing for the measurement of biologically meaningful signal differences between images. Intensity normalized images exist on an arbitrary but constant intensity scale which we use in place of quantitative relaxometry [20]. Affine registration of bispectral histograms does not account for variability of *F* across voxels or subjects, which makes the utility of the present method an empirical question. The results shown in Figures 5, 6, and 8 – 10 suggest that the present method has utility in the extraction of quantitative tissue biomarkers as well as lesion segmentation.

Intensity normalization yields reproducible images. Intensity normalized histograms are nearly indistinguishable across sessions within subjects but significantly differ across subjects (Figure 5). We have previously reported the use of tissue mean intensity and variability as a biomarker of MS severity [3]. In the present results these quantities exhibit ICC values above 0.75 on the basis of WTA segmentation (NAWM, GM, Lesion; Figure 9; Table 1). Similarly, using an external subcortical tissue segmentation algorithm, repeatability of tissue mean intensity and variability remained high (ICC greater than 0.67, Table 2). Therefore, intensity normalization yields reliable intensity maps that are stable in a test-retest scheme.

Image segmentation, particularly WM lesion (WML) segmentation, is a well developed technique [4, 12, 30, 32]. Algorithms for automatic tissue segmentation use a variety of approaches including probabilistic mapping and machine learning (for review, see [19, 12]). Tissue segmentation tools (e.g., FAST, FreeSurfer [11, 14]) as well as many WML segmentation tools (e.g., using machine learning algorithms [4, 19]) use both tissue intensity and anatomical priors. Here, we demonstrate tissue segmentation with minimal dependence on anatomical priors. The use of a simple WM prior was needed to constrain the identification of WMLs owing to large factitious lesion scores at the brain edge. Similarly, tissue score gradients were used to improve lesion segmentation as shown in Figure 9. Thus, in addition to extraction of intensity-based biomarkers, intensity normalization has value in image segmentation.

Tissue score gradients describe the transition from low to high tissue class scores. Considered in pairs, these gradients describe the transition from one tissue class to another. In this way, tissue transitions that are unlikely can be identified and heuristically corrected. For example, in the present work, the transition from Lesion to GM near a lesion cluster is unlikely. In the present application, consideration of tissue score gradients facilitated correction of factitious labeling of GMs around lesions. We note that juxtacortical and GM lesions do occur in MS.

Lesion identification *per se* is only one application of image intensity gradients. In both MS and cerebral small vessel disease, WMLs are dynamic entities capable of expansion or contraction [8, 7] or histological conversion e.g., into black holes [25]. Consideration of tissue gradients potentially allows for the sub-voxel determination of lesion boundaries as described in Figure 1. In this way, given the established reliability of intensity normalization, the boundary of lesions can be assessed longitudinally. Similarly, the existence of tissue gradients within structures (e.g., lesions) may indicate differential tissue damage and may represent an approach to longitudinal monitoring more specific than atrophy. Potential applications of tissue gradients will be assessed in future work.

## Data Availability

Imaging data are available through data sharing agreements from the study sponsor. All code and analytic tools are available upon reasonable request to the authors.

## Acknowledgments

MS PATHS is funded by Biogen. Dr.Brier was supported by 2R25NS090978-06. Dr. Xiang was supported by the National MS Society FG-1908-34882. Dr. Cross was supported by the Manny & Rosalyn Rosenthal – Dr John L. Trotter MS Center Chair in Neuroimmunology. Dr. Snyder was supported by P30NS098577 and P50HD103525. Dr. Benzinger was supported by 1R01AG054567- 01A1, P01AG003991, and U19AG03243808.

## Footnotes

1 Here we use the original notation in [17]. For clarity, *I*(1:2) is equivalent to the more modern notation, *D*_*KL*_(*P*_1_(*T*1*w, FLAIR*)|*P*_2_(*T*1*w, FLAIR*)), where *P*_1_ and *P*_2_ represent intensity distributions coresponding to different tissue classes.

2 Assuming that MS disease pathophysiology did not progress measurably over 7 days, given no clinical relapses in the cohort.

3 http://4dfp.readthedocs.io

